# Study protocol: a randomized controlled trial evaluating the effectiveness of a novel in-situ forming bone graft for lateral bone augmentation in compromised alveolar sites

**DOI:** 10.1101/2025.06.19.25329962

**Authors:** Nasim Safdarian, Amir Jalal Abbasi, Amir Reza Rokn, Ahmad Reza Shamshiri, Hamid Mobedi

## Abstract

**Background:** Synthetic bone substitutes offer a promising alternative to autografts, allografts, and xenografts for bone tissue regeneration, particularly for lateral bone augmentation (LBA) in the alveolar ridge. Traditional guided bone regeneration techniques for horizontal and vertical ridge defects are complex and associated with higher complication risks. This study introduces a novel in-situ forming scaffold (IFS) that solidifies at the surgery site, providing a stable framework for bone growth. Unlike animal-derived grafts, the IFS scaffold minimizes immunogenicity and disease transmission risks. This trial marks the first-in-human clinical evaluation of a patented IFS following successful preclinical studies. Its safety and efficacy will be assessed in comparison to Bio-Oss Spongiosa through radiographic and histological analyses, offering a novel synthetic approach to bone regeneration.

**Methods:** This randomized, controlled clinical trial will include 37 participants with lateral alveolar bone defects in the anterior region. Participants will be randomized to receive either Bio-Oss Spongiosa or the IFS scaffold. The primary outcomes include the scaffold’s safety and effectiveness in promoting bone regeneration, assessed radiographically and histologically at baseline, postoperatively, and after a 7.5-month healing period. Secondary outcomes involve implant stability and bone formation at re-entry. Blinding will be maintained for participants, outcome assessors, and statisticians.

**Discussion:** This trial represents the first evaluation of an in-situ forming scaffold for LBA. The IFS scaffold is expected to provide outcomes comparable to traditional grafts in bone fill, stability, and implant support, offering a safer synthetic alternative for alveolar bone regeneration. If successful, the findings may significantly advance techniques for alveolar ridge augmentation.

## Introduction

### Background and rationale

Bone tissue regeneration remains a critical challenge in oral and maxillofacial surgery, particularly in cases of lateral bone augmentation (LBA) for the alveolar ridge. The restoration of horizontal and vertical bone defects is essential to achieve optimal implant placement and functionality [1, 2]. However, traditional approaches, such as guided bone regeneration (GBR), autografts, allografts, and xenografts, present significant limitations [1, 3].

Autografts are considered the gold standard for bone regeneration due to their osteoconductive, osteoinductive, and osteogenic properties. However, they are associated with donor site morbidity, limited availability, and prolonged surgical time [4]. Allografts and xenografts, while more readily available, carry risks of immunogenicity and disease transmission [5-8]. These factors drive the search for synthetic alternatives that can provide comparable or superior outcomes without these inherent drawbacks.

Synthetic bone substitutes, or scaffolds, have emerged as a promising alternative. These scaffolds provide a framework for cell attachment, proliferation, and differentiation, ultimately facilitating new bone formation [9]. Among these, Bio-Oss Spongiosa, a deproteinized bovine bone mineral, is widely used due to its favorable biological and mechanical properties [3]. However, as an animal-derived product, it still poses risks of disease transmission and may not be acceptable to all patients due to ethical or cultural concerns [4, 5].

To address these limitations, this study introduces a novel in-situ forming scaffold (IFS) designed to solidify directly at the surgical site. This innovative scaffold eliminates the risks associated with immunogenicity and disease transmission inherent to animal-derived grafts. Additionally, it simplifies the surgical procedure by providing a stable, adaptable framework for bone regeneration. The IFS scaffold’s unique formulation is designed to enhance biocompatibility, osteoconductivity, and structural stability, offering a synthetic alternative that may overcome the challenges associated with traditional grafting materials [10].

Preclinical studies on the IFS scaffold have demonstrated its potential to support bone formation and maintain stability under various physiological conditions. Initial evaluations have shown promising results in terms of biocompatibility and integration with native bone tissue [10]. However, no clinical trials have yet assessed its safety and efficacy in humans. This trial aims to fill this gap by comparing the IFS scaffold to Bio-Oss Spongiosa through rigorous radiographic and histological assessments.

Lateral bone augmentation is critical for patients with compromised alveolar ridges, where insufficient bone volume limits the success of dental implants [2]. Current approaches using autografts, allografts, and xenografts have inherent limitations, as discussed. The IFS scaffold represents a potentially transformative solution, offering a synthetic, user-friendly alternative that minimizes risks while maintaining efficacy [10].

By conducting a randomized, controlled clinical trial, this study aims to generate high-quality evidence on the performance of the IFS scaffold. The comparison with Bio-Oss Spongiosa, a widely accepted standard in bone grafting [11, 12], will provide valuable insights into its relative benefits and limitations. The results of this trial could pave the way for broader adoption of synthetic scaffolds in clinical practice, reducing reliance on animal-derived materials and advancing the field of alveolar bone augmentation.

### Objetives

#### Primary objective

To evaluate the safety and efficacy of the in-situ forming scaffold (IFS) for lateral bone augmentation (LBA) in buccal bone defects compared to Bio-Oss Spongiosa, as assessed through radiographic measurements of bone width changes and histological evaluations. This will establish the minimum acceptable efficacy required for progression to phase II clinical trials, contingent on the absence of adverse effects and the demonstration of safety.

#### Secondary objectives

1. To assess the ability of the IFS scaffold to promote implant stability through resonance frequency analysis (RFA) at re-entry [13].
2. To evaluate the extent and quality of bone formation at re-entry, as measured by histomorphometric analyses, including parameters such as new bone formation, residual graft material, and total mineralized tissue [14].
3. To document the clinical parameters of soft tissue health, including probing pocket depth (PPD), bleeding on probing (BOP), and keratinized mucosa (KM) width at baseline and follow-up [15].
4. To compare patient-reported outcomes related to functionality, pain, and quality of life using validated scales, such as the Oral Health Impact Profile (OHIP-14) [16].
5. To monitor and document adverse events, postoperative complications, and overall safety profiles associated with the use of IFS scaffold and Bio-Oss Spongiosa [17, 18].

#### Exploratory objectives

1. To analyze the impact of IFS scaffold on the long-term dimensional stability of regenerated bone after a 12-month follow-up [19].
2. To investigate the potential of the IFS scaffold for simultaneous vertical and horizontal bone augmentation in complex defect cases.

These objectives are supported by previous findings highlighting the promise of synthetic scaffolds in bone regeneration while addressing limitations associated with traditional grafting materials.

#### Trial design

This randomized, controlled, parallel-group clinical trial aims to evaluate the safety and efficacy of an in-situ forming scaffold (IFS) versus Bio-Oss Spongiosa for lateral bone augmentation (LBA) in buccal bone defects. Participants will be randomized in a 1:1 ratio to receive either the IFS scaffold (intervention group) or Bio-Oss Spongiosa (control group). The study is designed to assess the superiority of the IFS scaffold in terms of radiographic bone width gain, histological outcomes, safety, and patient-reported measures.

## Methods: participants, interventions and outcomes

### Study setting

This study will be conducted at the Centre of Implantology, Dentistry Faculty, Tehran University of Medical Sciences, a leading academic and clinical institution for dental research and treatment. The study will take place exclusively in Iran, and additional details on study sites can be obtained directly from the corresponding research team or the trial registry entry.

#### Eligibility criteria

##### Inclusion Criteria

1. Adults aged 18 to 65 years.
2. Patients requiring lateral bone augmentation (LBA) in the anterior maxilla or mandible for dental implant placement.
3. Adequate vertical bone height but a horizontal bone width of ≤4 mm at the alveolar crest, confirmed by cone-beam computed tomography (CBCT).
4. Good general health, defined as no systemic diseases that could interfere with bone healing (e.g., uncontrolled diabetes, osteoporosis).
5. Good oral hygiene, assessed by a plaque index score of <1 and no active periodontal disease.
6. Willingness to adhere to study protocols and attend all follow-up visits.
7. Ability to provide written informed consent.

##### Exclusion Criteria

1. Systemic conditions known to affect bone metabolism or healing, such as osteoporosis, uncontrolled diabetes, or malignancies.
2. Current or recent use of medications affecting bone metabolism (e.g., bisphosphonates, corticosteroids).
3. Active oral or systemic infections.
4. History of smoking within the last 12 months.
5. Previous bone augmentation procedures at the intended implant site.
6. Pregnant or breastfeeding women.
7. Known allergies to any components of the scaffolds or membranes used in the study.
8. Participation in another clinical trial within the past six months.

#### Eligibility criteria for personnel

##### Personnel

Interventions will be performed by certified oral surgeons or periodontists with a minimum of five years of clinical experience in guided bone regeneration and implant placement. Training sessions will ensure consistent application of surgical protocols across all personnel.

These eligibility criteria ensure the selection of participants who are representative of the target population while minimizing risks and variability in outcomes.

#### Who will take informed consent?

Informed consent will be obtained by trained members of the research team, including the principal investigator, study coordinators, and dental specialists with clinical trial experience and familiarity with the study protocol. The process will begin with an information session, during which study objectives, procedures, risks, and benefits will be explained using simple, non-technical language. Participants will receive a written informed consent form (ICF) that complies with local ethical guidelines, institutional policies, and the Declaration of Helsinki.

Participants will have an opportunity to ask questions before signing the ICF. A witness will co-sign the form if required by institutional regulations. The signed forms will be securely stored, and participants will receive a copy for their records. In cases involving vulnerable individuals, authorized surrogates may provide consent. Consent forms will be available in the participant’s native language, and assistance will be provided for those with limited literacy.

This structured and ethically sound process ensures that all participants are fully informed and provide voluntary consent prior to enrollment in the trial.

#### Additional consent provisions for collection and use of participant data and biological specimens

Participants will be asked to provide optional, additional consent for the collection, storage, and future use of their clinical data and biological specimens in ancillary studies beyond the primary objectives of this trial. This includes clinical, demographic, and radiographic data, as well as biopsy samples collected during re-entry procedures. These materials may be used in future histological, molecular, or genetic analyses related to bone regeneration and scaffold performance.

The process will involve a separate, clearly distinguished consent form, accompanied by a detailed explanation of how the data and specimens may be used and how participant confidentiality will be protected. Participants may decline or withdraw this additional consent at any time without affecting their continued participation or care within the main trial.

By obtaining additional consent, this trial ensures ethical use of participant data and biological specimens, adhering to SPIRIT guidelines and the Declaration of Helsinki.

### Interventions

#### Explanation for the choice of comparators

The comparator selected for this study is **Bio-Oss Spongiosa**, a deproteinized bovine bone mineral that has been extensively used in lateral bone augmentation (LBA) and other guided bone regeneration (GBR) procedures. Bio-Oss Spongiosa is widely considered the gold standard for xenogeneic graft materials due to its established clinical success, osteoconductive properties. Its selection as the comparator provides a robust benchmark against which the performance of the investigational in-situ forming scaffold (IFS) can be evaluated.

#### Intervention description

Participants will be randomly assigned to one of two groups for lateral bone augmentation (LBA) using guided bone regeneration (GBR).

Group 1 – Test Group (IFS Scaffold):

Participants will receive an in-situ forming scaffold (IFS) composed of polylactic acid (PLA) and tricalcium phosphate (TCP). The scaffold is applied directly to the buccal bone defect and solidifies within two minutes, forming a porous structure that supports new bone growth. A resorbable collagen membrane will be placed over the scaffold to maintain stability and prevent soft tissue infiltration. The material will be administered during the initial surgical procedure, followed by standard postoperative care with antibiotics and analgesics. Group 2 – Control Group (Bio-Oss Spongiosa):

Participants will receive Bio-Oss Spongiosa, a well-established bovine-derived xenograft with proven osteoconductive properties. The particles will be hydrated with saline and placed onto the defect site, followed by coverage with a resorbable collagen membrane. The graft will be applied during the initial surgery, with postoperative care consistent with clinical guidelines.

Both interventions are designed to support bone regeneration and will be performed under standardized surgical protocols.

#### Criteria for discontinuing or modifying allocated interventions

To protect participant’s safety and maintain study integrity, predefined criteria will guide decisions on discontinuing or modifying assigned interventions. Interventions may be discontinued in cases of serious adverse events such as graft exposure, infection, or systemic complications that cannot be managed with standard care [17, 18]. Discontinuation may also occur due to disease progression, site incompatibility, or the discovery of conditions that make continued participation unsafe [20, 21]. Participants may also withdraw voluntarily, or discontinuation may result from significant protocol violations.

Modifications to interventions may be made if localized infections are manageable with antibiotics, or if supplementary procedures (e.g., additional grafting) are needed to ensure bone regeneration [18]. Adjustments in care may also be made in response to newly diagnosed medical conditions, without removing participants from the trial [20-22].

All decisions will be reviewed by the Data Safety and Monitoring Committee (DSMC), documented in case report forms, and reported to the ethics board. Participants will be informed of any changes and provided with appropriate follow-up care. These procedures ensure ethical oversight and preserve the scientific validity of the study.

#### Strategies to improve adherence to interventions

To ensure participants follow the intervention protocols and contribute to the reliability of study outcomes, several adherence strategies will be implemented. Participants will receive preoperative education, including counseling and informational materials, to reinforce the importance of following postoperative instructions and attending scheduled follow-ups. Standardized postoperative care plans—including guidance on oral hygiene, diet, and medication—will be customized to individual needs.

Adherence will be monitored through participant self-reports, clinical evaluations, and pharmacy records. Scheduled follow-up visits at key intervals (1 week, 1 month, 3 months, and 7.5 months) will include assessments of healing and protocol compliance, supported by automated reminders. Incentives such as travel reimbursement or certificates of participation may be provided to encourage consistent engagement.

Participants will have access to a study coordinator for support and updates throughout the trial. All adherence-related data will be documented in case report forms (CRFs), and any non-compliance will be reviewed by the monitoring committee. Digital tools, such as mobile apps or online portals, may also be used to enhance participant engagement and track adherence in real time. These strategies are designed to promote protocol fidelity and maintain data integrity.

#### Relevant concomitant care permitted or prohibited during the trial

To reduce confounding and preserve internal validity, the trial outlines specific guidelines regarding permitted and prohibited concomitant treatments. Medications and interventions that may interfere with bone regeneration—such as bisphosphonates, denosumab, corticosteroids, immunosuppressants, adjunctive procedures (e.g., PRP, laser therapy), or any unapproved investigational products—are not allowed during the study [20, 21].

Permitted care includes standard antibiotics and analgesics (e.g., amoxicillin, ibuprofen), routine oral hygiene practices that do not disturb the surgical site, and medications for stable chronic conditions that do not affect bone metabolism. Emergency care unrelated to the intervention is also permitted and documented [17].

All concomitant treatments and procedures will be recorded in the case report forms (CRFs) [23]. Any use of prohibited treatments must be reviewed and approved by the principal investigator and the Data Monitoring Committee (DMC) [24]. If such a treatment is deemed medically necessary, the participant may be withdrawn from the intervention arm but will continue follow-up for safety and outcome analysis under the intention-to-treat principle.

#### Provisions for post-trial care

Participants in this trial will receive structured post-trial support to ensure their ongoing well-being. A follow-up appointment will be offered at 12 months post-surgery to assess long-term outcomes, including graft stability. Any complications identified during this visit will be treated free of charge by the clinical team.

Participants who received the in-situ forming scaffold (IFS) will continue to have access to follow-up care related to the intervention. If trial-related adverse events occur—such as graft failure or infections— comprehensive medical care, including procedures, medication, or hospitalization, will be provided at no cost [18]. Rehabilitation services, such as physical therapy or dietary counseling, may also be offered when appropriate.

The trial is covered by clinical trial insurance to compensate participants for any harm directly caused by their involvement. All adverse events will be reviewed by the Data Safety and Monitoring Committee (DSMC) to ensure proper response and participant protection.

Participants will be assigned a contact person for post-trial communication and support. Upon study completion, they will receive a summary of trial findings and any relevant health implications. These provisions meet SPIRIT guidelines and reflect the trial’s ethical responsibility to its participants.

#### Outcomes

The **primary outcome** of this trial is the change in lateral bone width at the augmented site, assessed using cone-beam computed tomography (CBCT). Scans will be obtained preoperatively, immediately after grafting, and at 7 months postoperatively. The final measurement—taken approximately two weeks before implant placement—is used to calculate the change in bone width. A minimum gain of 3 mm (from a baseline ridge width of ≤4 mm) is considered a successful outcome. Volumetric assessments will also be performed using ImageJ software, applying Uchida’s method to estimate graft volume [25]. These imaging outcomes reflect the clinical effectiveness of bone augmentation for implant preparation [26].

In addition, histological analysis will be conducted on bone biopsies collected during the re-entry procedure at 7.5 months. Non-decalcified sections will be analyzed for proportions of new bone, residual graft, and connective tissue. New bone formation of >20% in the defect region will be considered a successful regenerative outcome [26, 27].

**Secondary outcomes** include:

- Implant stability, assessed through resonance frequency analysis (RFA) using the Osstell device. An implant stability quotient (ISQ) value >60 will be considered indicative of successful osseointegration [13].
- Radiographic bone fill, calculated as the percentage of defect fill on CBCT scans from baseline to 7.5 months, providing an additional measure of bone regeneration [28].
- Patient-reported outcomes, evaluated using the OHIP-14 questionnaire to measure changes in oral health-related quality of life from baseline to 7.5 months.
- Need for secondary augmentation, measured as the proportion of patients requiring additional grafting procedures in either group.

**Safety outcomes** include the incidence and severity of adverse events, which will be monitored continuously throughout the study to evaluate the safety profiles of both interventions.

#### Participant timeline

Participants in this study will follow a structured timeline from screening through extended follow-up to enable consistent evaluation of the intervention’s safety and efficacy [figure 1]. During the screening phase (weeks - 2 to 0), eligibility will be confirmed, and baseline clinical and radiographic assessments will be completed. On the day of surgery (Day 0), participants will be randomized to receive either the in-situ forming scaffold (IFS) or Bio-Oss Spongiosa, followed by the lateral bone augmentation procedure using guided bone regeneration (GBR).

**Figure 1.**
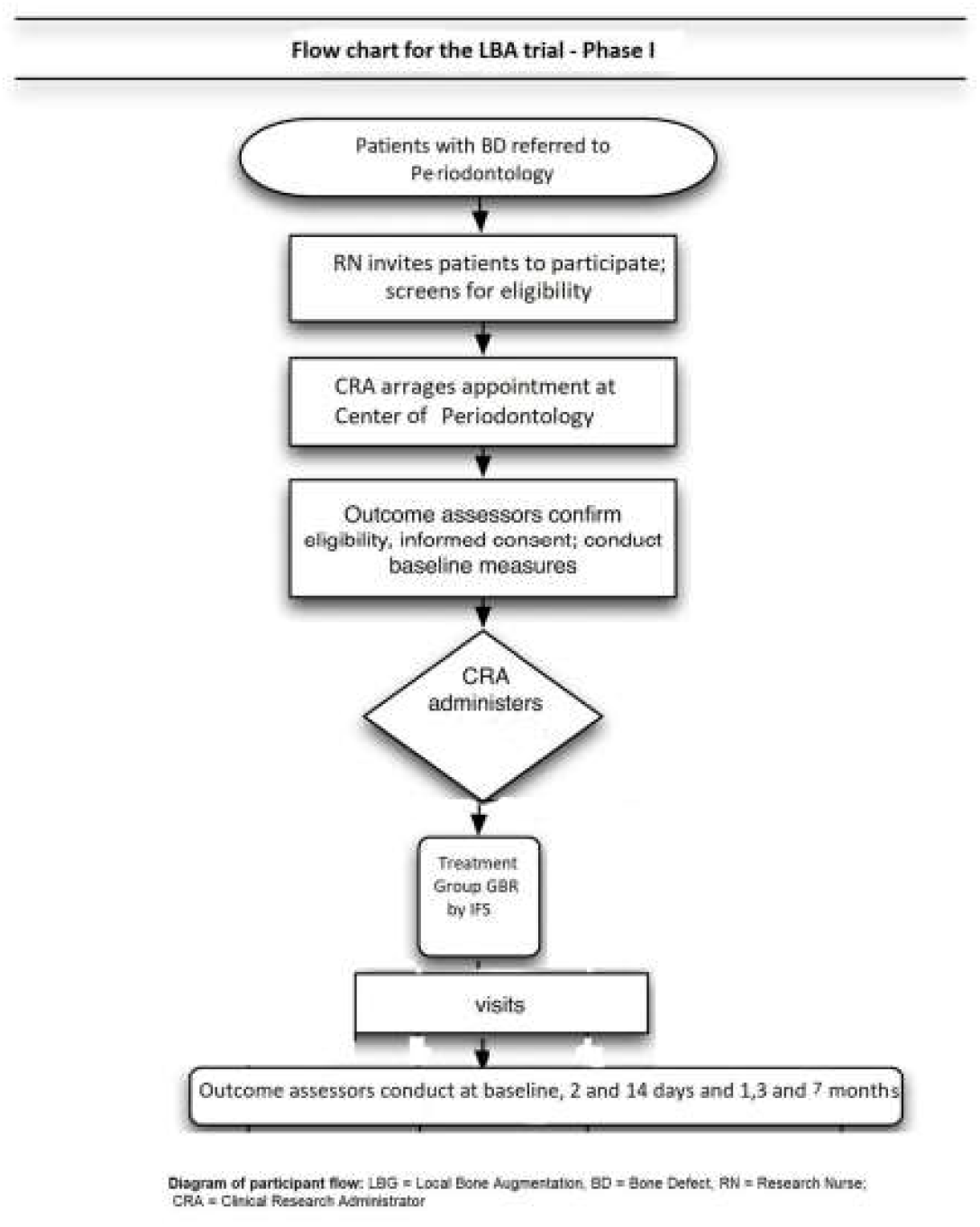
Participant Flow Chart for the LBA Trial – Phase I. *Diagram of participant flow. LBA = Local Bone Augmentation; BD = Bone Defect; RN = Research Nurse; CRA = Clinical Research Administrator*.

Postoperative follow-up visits will occur on Day 2 and Day 14 to assess wound healing and adverse events, and again at 1 and 3 months to monitor early outcomes. During the healing period (Months 4–7), participants will undergo additional assessments to evaluate graft stability. At Month 7, radiographic evaluation using CBCT and histological analysis of biopsies will be performed to assess bone regeneration. At Month 7.5, dental implants will be placed, and implant stability will be measured via resonance frequency analysis (RFA).

Participants will be followed for a total of 12 months to assess long-term outcomes, including the dimensional stability of the regenerated bone. Outcome assessments will be conducted by independent clinical, radiographic, and histological evaluators. The full timeline is presented in Table 1.

**Table 1.** Participant Timeline and Key Assessments.

| Phase | Time Frame | Activities/Assessments |
| --- | --- | --- |
| Screening Phase | Week -2 to 0 | Participant recruitment, eligibility screening, and baseline clinical and radiographic assessments. |
| Day of Surgery | Day 0 | Randomization and intervention (LBA using IFS or Bio-Oss Spongiosa with GBR). |
| Postoperative Follow-Up | Day 2 and Day 14 | Clinical exams to assess wound healing and monitor for adverse events. |
| Postoperative Follow-Up | Month 1 and Month 3 | Monitoring clinical healing and early outcomes. |
| Healing Period | Months 4–7 | Assessments as needed to evaluate graft stability and integration. |
| Primary Assessment | Month 7 | CBCT scans for bone width measurement and biopsies for histological evaluation. |
| Implant Placement | Month 7.5 | Placement of dental implants and RFA to assess implant stability. |
| Extended Follow-Up | Up to Month 12 | Monitoring long-term dimensional stability of regenerated bone. |

#### Sample size

A total of 37 participants will be enrolled in this trial to ensure sufficient power to detect a clinically meaningful difference in lateral bone width between the intervention and control groups. The sample size calculation was based on an expected effect size of 1.5 mm and a standard deviation of 1.0 mm, with a power of 80% and a two-sided significance level of 0.05 [29]. Using these parameters and the standard formula for comparing two means [30, 31], 15 participants per group were required. To account for an estimated 20% dropout rate, the final sample size was increased to 37.

Participants will be randomized in a 1:1 ratio into two groups: the intervention group (IFS scaffold, n = 18) and the control group (Bio-Oss Spongiosa, n = 19). This allocation ensures balanced representation and allows for valid statistical comparisons between groups.

#### Recruitment

To recruit the target sample of 37 participants, a comprehensive, multi-site recruitment strategy will be implemented [24, 25]. Participants will be enrolled through the Dental Faculty Clinic at Tehran University of Medical Sciences as well as affiliated partner clinics and private dental offices. Collaboration with regional specialists, including oral surgeons and periodontists, will support referrals, with informational sessions organized to increase clinician awareness and engagement.

Outreach strategies will include posters and flyers placed in clinic waiting areas, digital advertisements on institutional websites and social media, and community awareness events focused on bone augmentation. Interested individuals will be screened for eligibility and, if qualified, will be fully informed about the study before providing written consent.

To incentivize participation, individuals will receive free treatment—including lateral bone augmentation and implant placement—and may be reimbursed for travel or offered modest compensation for attending follow-up visits.

Recruitment will be closely monitored through weekly progress reports. If enrollment targets are not met, additional recruitment centers may be added and outreach efforts intensified to keep the study on schedule [24, 25].

### Assignment of interventions: allocation

#### Sequence generation

To ensure unbiased participant allocation, a computer-generated randomization sequence will be created using statistical software such as SPSS or R, assigning participants in a 1:1 ratio to the intervention group (IFS scaffold) or the control group (Bio-Oss Spongiosa) [24, 25].

A permuted block randomization method will be used with a block size of four to maintain group size balance. Randomized sequences within each block will reduce the predictability of assignment. Stratification will be applied based on baseline variables—age, sex, and defect size—to control for their potential influence on bone healing and ensure comparability across groups.

Allocation concealment will be maintained by an independent statistician, who will generate and store the randomization sequence in a password-protected file. Investigators and enrolling personnel will be blinded to group assignments [25].

Once eligibility is confirmed, the study coordinator will retrieve the allocation code from the statistician and relay it to the clinical team, ensuring that outcome assessors remain blinded [24, 25]. This allocation approach—using blocking, stratification, and concealment—strengthens internal validity and limits selection bias [25].

#### Concealment mechanism

To minimize selection bias and maintain allocation integrity, this study will use both centralized and backup allocation concealment strategies. An independent statistician—unaffiliated with recruitment or clinical procedures—will generate and store the randomization sequence in a password-protected database. Once a participant is deemed eligible and has consented, the study coordinator will contact the statistician to obtain the group assignment [24, 25].

As a secondary method, sequentially numbered, opaque, sealed envelopes containing preassigned group allocations will be prepared and securely stored. These will be opened only after participant enrollment is complete.

Blinding will be maintained throughout the trial. Participants will remain unaware of their assigned group; during surgery, graft application will occur without visual or verbal disclosure. The re-entry procedure will be conducted by a different surgeon to ensure unbiased outcome evaluation. Clinical, radiographic, and pathological outcome assessors—as well as data analysts—will remain blinded to group allocation. These procedures ensure robust allocation concealment and uphold the SPIRIT guidelines for methodological rigor [24, 25].

#### Implementation

The randomization sequence will be created using a stratified, permuted block design in SPSS by an independent statistician who is not involved in recruitment, intervention, or outcome assessment. The allocation list will be securely stored in a password-protected electronic database [24, 25].

Participant screening and enrollment will be managed by study coordinators at the Dental Faculty Clinic of Tehran University of Medical Sciences. Coordinators will confirm eligibility, obtain written informed consent, and remain blinded to the allocation sequence.

Following enrollment, the coordinator will contact the statistician to receive the group assignment. If the centralized system is unavailable, group allocation will be retrieved from a secure backup set of sequentially numbered, opaque, sealed envelopes. The assigned group (IFS scaffold or Bio-Oss Spongiosa) will then be communicated to the clinical team for intervention delivery.

Blinding will be maintained throughout the study. Participants, outcome assessors (clinical, radiographic, histological), and statisticians will remain blinded to group assignments. Additionally, the surgeon conducting the re-entry procedure will differ from the initial surgeon to reduce bias. Blinding measures, including sealed envelopes and coded labeling, will help prevent detection and performance bias [25].

This structure ensures secure and unbiased allocation while maintaining the scientific integrity of the trial.

### Assignment of interventions: blinding

#### Who will be blinded

To minimize selection and assessment bias, this trial will implement blinding for key stakeholders including participants, outcome assessors, and data analysts [24, 25]. Participants will remain unaware of their assigned intervention group. To maintain the blind, both the IFS scaffold and Bio-Oss Spongiosa will be prepared and applied in a standardized way to prevent visible or procedural differences. This approach ensures that participants’ perceptions and responses are not influenced by knowledge of the treatment received.

Outcome assessors—including clinicians, radiologists, and histologists—will be blinded to group assignments when performing clinical evaluations, radiographic measurements, and histological analyses. Data analysts will work with de-identified datasets coded by group, allowing for unbiased statistical interpretation [24, 25]. The re-entry surgeon will be different from the initial surgeon to further maintain the integrity of blinded assessments.

Care providers performing the surgical interventions and study coordinators responsible for enrollment and randomization will not be blinded, due to the practical requirements of clinical care and group assignment. However, their roles are limited to procedural duties and do not involve outcome measurement or data interpretation. Allocation concealment will be maintained using a password-protected list managed by an independent statistician. These measures align with SPIRIT guidelines and are designed to protect the objectivity and reliability of the trial’s findings [24, 25].

#### Procedure for unblinding if needed

Unblinding will only occur under exceptional circumstances when participant safety is at risk, such as in cases of serious adverse events (SAEs) like severe infection or unexpected healing complications requiring knowledge of the assigned graft material [24].

Formal requests for unblinding must be submitted by the treating clinician and reviewed by the principal investigator in consultation with the safety monitoring team. If approved, the study coordinator will obtain the allocation from the independent statistician or sealed envelope. The information will be shared only with essential clinical personnel, while efforts will be made to keep participants and outcome assessors blinded unless necessary [24, 25].

All unblinding events will be documented, including the rationale, timing, and involved personnel. To minimize bias, unblinded participants will be assessed by blinded evaluators when feasible, and data analysts will remain blinded to other participants’ group assignments. This protocol upholds ethical and scientific standards in line with SPIRIT guidelines [25].

### Data collection and management

#### Plans for assessment and collection of outcomes

This study uses standardized procedures and validated tools to ensure consistent and accurate outcome assessment across defined time points [32]. At baseline, participants will undergo clinical evaluation, CBCT imaging, and medical history documentation using a structured questionnaire. Postoperative assessments will occur on Day 2, Day 14, and at 1 and 7.5 months. At 7.5 months, CBCT will evaluate ridge width, and biopsies will be collected during re-entry for histological analysis [28].

Key instruments include CBCT for bone width measurement, histological analysis for bone quality, and resonance frequency analysis (RFA) to assess implant stability [13]. Radiologists and histologists will be calibrated, and duplicate readings will be used to ensure data reliability [32].

All data will be entered into Good Clinical Practice (GCP)-compliant, password-protected electronic case report forms (eCRFs), with access granted through the trial registry upon request. Regular internal audits and oversight by an independent monitoring committee will ensure protocol adherence and ethical compliance. This approach ensures robust data quality to evaluate the scaffold’s safety and effectiveness.

#### Plans to promote participant retention and complete follow-up

To reduce dropout and ensure complete follow-up, this study applies structured retention strategies that prioritize participant support while maintaining protocol adherence [25]. Participants will receive regular reminders via phone, email, or SMS, and study coordinators will maintain personalized communication throughout the trial.

Flexible scheduling, including evening or weekend appointments, will be offered to accommodate participant needs. Travel reimbursements and modest incentives will also be provided. Participants will be kept engaged through periodic updates about the study’s progress and transparent communication about data use and confidentiality [24].

For participants who discontinue or deviate from the intervention, efforts will be made to collect partial outcome data such as final CBCT scans [28], histological samples (when consented), and patient-reported outcomes. To address missing data, statistical methods like multiple imputation and sensitivity analyses will be applied in line with current best practices [32].

Retention will be monitored continuously. Early signs of disengagement will prompt intervention, including additional support or counseling, to maintain participant involvement and study integrity [24, 25].

#### Data management

This study follows a robust data management plan to ensure data accuracy, security, and regulatory compliance [32]. All participant data will be entered into a secure, password-protected electronic data capture (EDC) system. Critical outcomes, such as radiographic and histological data, will undergo double data entry by trained staff, with discrepancies resolved by a third reviewer.

Participants will be assigned unique alphanumeric codes to ensure anonymity. Clinical variables and adverse events will be coded using standardized formats, including ICD-10 classifications when applicable. The EDC system includes built-in range and consistency checks, with audit trails to track any data changes.

Access to the database will be restricted to authorized personnel, and all data will be encrypted during storage and transmission. Regular backups will be performed, and both electronic and physical records will be stored securely. Data will be archived for at least 10 years following publication.

Further details, including coding systems, error handling workflows, and the data dictionary, are outlined in the study’s Data Management Plan (DMP), available upon request from the sponsor [32].

#### Confidentiality

To protect participant confidentiality and comply with Good Clinical Practice (GCP) and ethical guidelines, several measures will be enforced throughout the trial [24, 25]. Personal data, including contact details and medical history, will be collected during screening and enrollment. Each participant will be assigned a unique alphanumeric code for all future data collection to ensure anonymity.

Only de-identified data will be shared with outcome assessors, data analysts, and external collaborators. Access to identifiable information will be restricted to authorized staff such as the principal investigator and study coordinators. Paper documents will be stored in locked cabinets, while electronic data will be kept in password-protected, encrypted databases with audit trails and regular server backups [32].

During the trial, coded identifiers will replace personal details in all communications. Data transfers, including those for monitoring or statistical analysis, will be encrypted. Personal data will be stored for at least 10 years after publication, remaining anonymized and excluded from any published results or presentations [25].

The study’s confidentiality procedures will be reviewed by the institutional ethics committee before trial initiation. Compliance will be monitored throughout the study by the sponsor and an independent monitoring committee to ensure adherence to all applicable standards [25].

#### Plans for collection, laboratory evaluation and storage of biological specimens for genetic or molecular analysis in this trial/future use

N/A

In this trial, blood and urine samples will be collected for safety evaluation of the investigational product. Participants will be referred directly to the laboratory for sample collection prior to surgery and at two post-operative time points (2 days and 1 month after surgery). These samples will be analyzed promptly for safety assessments. No biological specimens will be stored for future genetic or molecular analysis, and no biobanking is planned.

### Statistical methods

#### Statistical methods for primary and secondary outcomes

This study will apply robust statistical techniques to evaluate primary and secondary outcomes, following a detailed Statistical Analysis Plan (SAP) available upon request [25].

The primary outcome—change in lateral bone width from baseline to 7.5 months—will be analyzed using paired t-tests or Wilcoxon signed-rank tests for within-group comparisons, and independent t-tests or Mann-Whitney U tests for between-group comparisons, depending on data normality [25, 30, 31]. Covariates such as age, sex, and defect size will be adjusted using a general linear model (GLM).

For secondary outcomes, implant stability (measured by RFA) will be analyzed with repeated-measures ANOVA [13]. Histological data, including new bone and residual graft percentages, will be summarized descriptively and compared using t-tests or non-parametric equivalents if needed [27]. Soft tissue health parameters (e.g., probing depth, bleeding, keratinized mucosa width) will be analyzed using mixed-effects models to account for repeated measures [33].

Missing data will be addressed using multiple imputation or sensitivity analyses. Both intention-to-treat (ITT) and per-protocol (PP) analyses will be conducted to ensure comprehensive interpretation [32].

All analyses will be performed using SPSS v28.0 or R v4.2. An independent statistician will validate the code and results to ensure accuracy and reproducibility [30, 31].

#### Interim analyses

Interim analyses will be conducted to monitor safety, efficacy, and trial progress, ensuring ethical oversight and scientific validity [24, 25, 32]. A safety analysis will occur after approximately 30% of participants (around 18) have completed the 7.5-month follow-up. An interim efficacy analysis will also evaluate changes in lateral bone width to determine whether the trial should continue as planned or be modified or stopped early.

Stopping guidelines include early termination i0066 there is a significant increase in serious adverse events (SAEs), if a statistically significant benefit is observed using pre-defined thresholds (adjusted via the O’Brien-Fleming method), or if futility analysis suggests the study is unlikely to detect a meaningful difference between groups. An independent Data Monitoring Committee (DMC) will review interim results and advise on whether to continue, modify, or stop the trial. The DMC will have access to unblinded data, while investigators and sponsors will remain blinded to maintain objectivity [32]. Final decisions will be made by the principal investigator in consultation with the sponsor.

Statistical analysis for interim reviews will follow group sequential methods with Type I error control, using techniques such as O’Brien-Fleming or Pocock boundaries to preserve the integrity of the overall study design [30, 31].

#### Methods for additional analyses (e.g. subgroup analyses)

N/A

This study is designed as a Phase I/II clinical trial primarily focused on the safety and efficacy evaluation of a novel in-situ forming scaffold (IFS) for lateral bone augmentation. Given the early-stage, proof-of-concept nature of the trial and the limited sample size, subgroup or adjusted analyses (e.g., based on age, sex, or baseline defect size) are not planned.

#### Methods in analysis to handle protocol non-adherence and any statistical methods to handle missing data

This trial includes interim analyses to assess safety, efficacy, and overall progress, ensuring ethical and scientific oversight [24, 25, 32]. Safety data will be reviewed after 30% of participants complete the 7.5-month follow-up. Efficacy will also be assessed to determine if early trial termination is warranted.

Stopping decisions may be based on significant safety concerns, early evidence of benefit (using O’Brien-Fleming thresholds), or futility. An independent Data Monitoring Committee (DMC) will evaluate unblinded interim data, while investigators and sponsors remain blinded. Recommendations from the DMC will guide final decisions by the principal investigator and sponsor [32].

Statistical methods will use group sequential designs with adjustments (e.g., O’Brien-Fleming or Pocock) to control Type I error and maintain study integrity [30, 31].

#### Plans to give access to the full protocol, participant level-data and statistical code

The full trial protocol will be publicly accessible following publication of the primary results. It will be available via the Iranian Registry of Clinical Trials (IRCT) and included as supplementary material in related peer-reviewed articles [24, 25].

De-identified participant-level data will be shared with qualified researchers upon request, pending ethical approval and submission of a clear research plan. Access will be granted after publication of the main findings and hosted on secure repositories such as Dryad, Figshare, or institutional platforms [24, 25].

The statistical code used for data analysis will also be made publicly available in open-access repositories like GitHub or Zenodo under an open-source license (e.g., GNU GPL) to support transparency and reproducibility [24, 32].

Access to both data and code will be governed by a Data Use Agreement (DUA) to ensure ethical, legal, and regulatory compliance, including adherence to GDPR and other applicable data protection standards [25, 32].

### Oversight and monitoring

#### Composition of the coordinating center and trial steering committee

The trial will be overseen by multiple independent committees to ensure compliance with Good Clinical Practice (GCP) and to maintain ethical and scientific integrity throughout the study [24, 25].

The Coordinating Centre at Tehran University of Medical Sciences will manage day-to-day operations, including participant coordination, data management, and communication between sites. This team, led by the principal investigator, will meet biweekly.

A Trial Steering Committee (TSC)—comprising independent clinicians, statisticians, sponsor representatives, and an ethics advisor—will oversee the overall conduct of the study, review protocol amendments, and ensure adherence to regulatory requirements. The TSC will meet quarterly or as needed.

An Endpoint Adjudication Committee (EAC) of independent experts in radiology and histology will validate outcome data at key timepoints, including the 7.5-month assessment.

The Data Monitoring Committee (DMC), consisting of independent biostatisticians and clinicians, will review safety data and conduct interim analyses. Meetings will occur at predefined interim checkpoints or if safety concerns arise.

This oversight structure provides comprehensive monitoring at both operational and strategic levels, ensuring data integrity, participant safety, and regulatory compliance throughout the trial [Table 2].

**Table 2:** Trial Oversight Structure and Committee Scheduling.

| Committee | Frequency of Meetings |
| --- | --- |
| Coordinating Centre | Biweekly |
| Trial Steering Committee | Quarterly and as needed |
| Endpoint Adjudication Committee | Post-data collection milestones |
| Data Monitoring Committee | Interim analysis and as needed |

#### Composition of the data monitoring committee, its role and reporting structure

##### Data monitoring committee (DMC)

The DMC is composed of five independent experts—two clinical specialists in bone regeneration, two biostatisticians, and one ethics advisor—to ensure objective oversight. All members operate independently of the sponsor (NIMAD) and have signed conflict of interest declarations [24, 25].

The committee is responsible for monitoring participant safety by reviewing adverse events (AEs and SAEs), evaluating interim efficacy data, and ensuring ethical compliance throughout the trial [24, 25]. Recommendations from the DMC are reported to the Trial Steering Committee (TSC) and shared with the coordinating center for action once approved.

DMC meetings are scheduled: Prior to trial initiation, after interim analyses, upon the occurrence of safety concerns, and at study completion. A formal DMC charter outlines roles, meeting procedures, and reporting protocols. This committee is essential given the surgical nature of the trial and the potential risks associated with bone augmentation [24, 25].

#### Adverse event reporting and harms

##### Adverse event reporting and management

This trial follows Good Clinical Practice (GCP) guidelines and international standards for collecting, assessing, reporting, and managing adverse events (AEs) to ensure participant safety and trial integrity [18, 22, 24, 25]. AEs will be gathered systematically during follow-up visits (Days 2 and 14, Months 1, 3, and 7.5) using structured questionnaires, as well as through 24/7 participant reporting. All events—including onset, severity, duration, and suspected cause—will be recorded in electronic case report forms (eCRFs). In guided bone regeneration (GBR) procedures, membrane exposure is a known risk that can compromise outcomes, particularly in esthetic areas. Other risks include infection and abscess formation, even without membrane exposure [17, 18].

Events will be graded using the Common Terminology Criteria for Adverse Events (CTCAE) scale and categorized by causality. GBR-specific classification systems, such as Fontana’s membrane exposure grading (Classes I–IV), and surgical complication types (e.g., flap damage, nerve injury) will be used for consistency [17, 18, 34].

Serious adverse events (SAEs), such as hospitalization or life-threatening conditions, must be reported within 24 hours to the DMC and regulatory authorities. Routine AEs will be summarized in monitoring reports and annual safety reviews [22]. AEs will be managed clinically, and protocol modifications may be made if recurring patterns emerge, pending IRB approval. Participants with severe or intolerable AEs may be withdrawn while retaining their data for analysis [17, 18, 22].

The DMC will conduct ongoing safety reviews and interim analyses, while the Trial Steering Committee will oversee protocol adherence [24, 25]. All AE records will be securely stored in the eCRF system with full audit trails to meet regulatory standards [23, 32].

#### Frequency and plans for auditing trial conduct

##### Auditing trial conduct

To ensure compliance with Good Clinical Practice (GCP), ethical standards, and regulatory requirements, independent audits will be conducted at key stages of the trial [24, 25]. These include pre-trial (to assess site readiness), at the midpoint (after 50% of participants are enrolled), and post-trial (after follow-up concludes). Additional audits may occur in response to safety concerns or protocol deviations, as recommended by the Data Monitoring Committee (DMC).

Audits will be carried out by an external team not involved in the trial. The scope includes reviewing recruitment practices, informed consent procedures, adverse event reporting, data handling, and overall protocol adherence. Findings will be reported to the Trial Steering Committee (TSC) and sponsor for action.

Auditors will operate independently of the sponsor and investigators, with oversight from the TSC to maintain objectivity and accountability [25]. Any deficiencies identified will be addressed promptly through corrective actions such as retraining, protocol amendments, or increased monitoring [24, 25].

#### Plans for communicating important protocol amendments to relevant parties (e.g. trial participants, ethical committees)

##### Protocol amendments

Protocol changes will be managed systematically to maintain ethical compliance and transparency. All amendments—triggered by new safety data, operational needs, or committee recommendations—will be documented with justification and impact details using formal amendment forms [25].

Prior to implementation, amendments will be submitted for approval to Institutional Review Boards (IRBs) or Ethical Review Committees (ERCs). In urgent situations affecting participant safety, immediate changes may be enacted and reported retrospectively. Major modifications will also be sent to regulatory authorities for approval [24, 35].

Participants impacted by changes (e.g., eligibility criteria or visit schedules) will be notified directly, and updated informed consent will be obtained if the risk-benefit profile is affected [25]. Investigators and study staff will receive formal protocol update training, with revised documents distributed and archived accordingly. Significant amendments will be updated in trial registries such as IRCT and, if the protocol is published, reflected in journals through addenda or errata [35]. The Trial Steering Committee (TSC) will monitor and approve the entire amendment process to ensure full compliance with ethical and regulatory standards [24, 25].

#### Dissemination plans

##### Dissemination plans

The results of this trial will be shared with participants, healthcare professionals, and the public in accordance with GCP and international reporting standards [25].

Participants will receive a lay summary of findings, including the outcome of their assigned group (IFS scaffold or Bio-Oss Spongiosa), delivered during follow-up or via written communication based on preference. Personal data will remain confidential and will not be disclosed [24, 25].

Healthcare professionals will be informed through presentations at national and international scientific conferences (e.g., IADR) and through educational workshops related to dentistry and bone regeneration.

Public dissemination will include plain-language summaries posted on the Iranian Registry of Clinical Trials (IRCT) and institutional websites [35]. Press releases will be shared with media outlets to enhance public awareness.

Scientific publications will include the main findings published in high-impact, peer-reviewed journals such as *Clinical Oral Implants Research* and *Journal of Advanced Prosthodontics*. Articles will be open access whenever possible. Secondary and exploratory findings will be shared through additional publications.

Results databases like IRCT and ClinicalTrials.gov will be updated with final outcomes [35]. De-identified participant-level data and statistical code will be available upon reasonable request, encouraging collaboration and secondary research [24, 25].

There are no restrictions from the sponsor (NIMAD) regarding publication, and authorship will follow recognized international guidelines to ensure appropriate contributor recognition.

## Discussion

This clinical trial addresses the significant need for advanced materials in bone regeneration, focusing on lateral bone augmentation (LBA) techniques. Current methods, while effective in some contexts, present limitations such as donor site morbidity in autografts, potential immunogenic responses in allografts, and ethical concerns surrounding xenografts [4-8]. The use of a novel in-situ forming scaffold (IFS) represents an innovative approach to overcoming these challenges.

A critical operational consideration in this trial is the implementation of rigorous randomization and blinding protocols to ensure unbiased results. Employing computerized randomization and stratification by patient characteristics, such as defect size and patient age, enhances the study’s internal validity. This level of methodological rigor is essential to minimize selection and measurement bias [24, 25].

From a logistical perspective, one anticipated challenge is ensuring participant adherence to the follow-up schedule, particularly given the extended monitoring period of 12 months. Strategies such as travel reimbursements and regular reminders have been incorporated to mitigate this risk and maintain participant retention [25].

Another significant aspect of this trial is the comprehensive collection of outcomes, including radiographic, histological, and patient-reported measures [16]. The incorporation of advanced imaging techniques, such as cone-beam computed tomography (CBCT), ensures precise evaluation of bone regeneration [28]. Meanwhile, the use of validated scales, like the Oral Health Impact Profile (OHIP-14), enables the assessment of functional and quality-of-life outcomes [24, 25].

However, this study is not without limitations. The single-center design may limit the generalizability of findings to other populations or clinical settings. Future multicenter trials could address this limitation by incorporating a broader participant base and diverse geographic regions [25].

The decision to use Bio-Oss Spongiosa as a comparator is supported by its widespread acceptance as a gold standard in bone grafting [11, 12, 36]. Nonetheless, the study’s exploratory objectives to evaluate IFS scaffold applications in vertical bone augmentation and complex defect cases highlight the potential for broader implications of the findings [10].

In conclusion, this trial aims to provide robust evidence on the safety and efficacy of the IFS scaffold in comparison to Bio-Oss Spongiosa. By addressing current limitations in bone augmentation techniques and exploring new frontiers in scaffold applications, this study has the potential to make a significant impact on clinical practices in dentistry and beyond.

### Trial status

The study received medical ethics approval from the Committee of the Health Minister of Iran in 2018. It took approximately six months to obtain funding, and patient recruitment began in late 2019. However, due to the outbreak of the COVID-19 pandemic, all trial activities were halted from late 2019 until late 2021. As a result, the trial has been postponed until now. Patient recruitment has resumed and is still ongoing. Recruitment started in 2024 and is expected to be completed around 2026.

## Data Availability

This manuscript describes the study protocol for a randomized controlled trial.

## Abbreviations

IRCT: Iranian Registry of Clinical Trials
IFS: In-situ Forming Scaffold
LBA: Lateral Bone Augmentation
GBR: Guided Bone Regeneration
RCT: Randomized Controlled Trial
CBCT: Cone-Beam Computed Tomography
RFA: Resonance Frequency Analysis
ISQ: Implant Stability Quotient
OHIP-14: Oral Health Impact Profile – 14-item
AE: Adverse Event
SAE: Serious Adverse Event
ITT: Intention-to-Treat
PP: Per-Protocol
PI: Principal Investigator
TSC: Trial Steering Committee
DMC: Data Monitoring Committee
TUMS: Tehran University of Medical Sciences
NIMAD: National Institute for Medical Research Development
CTCAE: Common Terminology Criteria for Adverse Events
SAP: Statistical Analysis Plan
DMP: Data Management Plan
DUA: Data Use Agreement
MSOP: Manual of Standard Operating Procedures
CRA: Clinical Research Administrator
RN: Research Nurse
OMLBACT: Outcome Measures in Lateral Bone Augmentation Clinical Trials
SF-12: Short Form 12-item General Health Questionnaire
BWS: Bone Width Scale
BD: Bone Defect

## Acknowledgements

We thank the National Institute for Medical Research Development (NIMAD) and Tehran University of Medical Sciences (TUMS) for providing the financial and institutional support that made this research possible. We extend our deepest gratitude to the participants who volunteered to take part in this research. Their trust and cooperation were fundamental to the success of this study.

## Clinical and Research Team

- Dr. Amirreza Rokn, Professor, Department of Dentistry, Tehran University of Medical Sciences, for his role as a Co-Investigator, supporting clinical procedures and participant follow-up.
- Dr. Amir Jalal Abbasi, Assistant Professor, Department of Dentistry, Tehran University of Medical Sciences, for serving as the Principal Investigator and leading the overall study design and execution.
- Dr. Hamid Mobedi, Chief Scientific Officer, EmerRx Biopharma company, for his contributions to scaffold material development and analysis.
- Dr. Ahmad Reza Shamshiri, Associate Professor, Department of Medical Epidemiology, Tehran University of Medical Sciences, for his expertise in statistical analysis and methodological design.
- Dr. Nasim Safdarian, Instructor, Departments of Orthodontics and Biomedical Sciences, Arthur A. Dugoni School of Dentistry, University of the Pacific, San Francisco, CA, USA., for her dedicated efforts as Project Manager, R&D researcher and ensuring effective coordination of all study activities.

## Authors’ contributions

The contributions of each author to this manuscript are outlined below, in accordance with international authorship guidelines:

- **AJA (Dr. Amir Jalal Abbasi):** AJA serves as the Principal Investigator. He conceives the study and leads the development of the research protocol. He provides oversight of study design and execution.
- **NS (Dr. Nasim Safdarian):** NS serves as the Project Manager. She coordinates the study, oversees participant recruitment, and manages logistics. She also contributes to manuscript preparation and critical review. Notably, NS is the R&D researcher of this product. The first inventor of US patent covering the novel in-situ forming scaffold evaluated in this trial..
- **AR (Dr. Amirreza Rokn):** AR contributes to clinical study design, performs surgical interventions, and supports participant follow-up. He reviews the manuscript with focus on clinical accuracy.
- **HM (Dr. Hamid Mobedi):** HM supports scaffold material development and advises on materials science aspects of the study. He contributes to manuscript review.
- **ARS (Dr. Ahmad Reza Shamshiri):** ARS designs the statistical methodology, analyzes the data, and writes the statistical analysis section of the manuscript.

## Availability of data and materials

The principal investigator, Dr. Amir Jalal Abbasi, will have full access to the study dataset to ensure scientific integrity. Co-investigators Drs. Safdarian, Rokn, Mobedi, and Shamshiri will have role-specific access related to clinical, statistical, or materials oversight. An anonymized dataset will be analyzed by a designated statistician under Dr. Shamshiri’s supervision.

There are no contractual limitations restricting data access. The funder, NIMAD, will not have access to or influence over data analysis or reporting.

De-identified data and statistical code may be shared with qualified external researchers upon approval by the Trial Steering Committee (TSC), pending submission of a research proposal and adherence to ethical standards [25, 32].

## Ethics approval and consent to participate

### Ethics Approval and Consent to Participate

The study was approved by the Ethics Committee of the National Institute for Medical Research Development (NIMAD), confirming compliance with national ethical standards.

- **Approval ID:** IR.NIMAD.REC.1397.303
- **Approval Date:** July 1, 2018

#### Approval Statement

The committee verified that the study meets ethical and legal requirements. The Principal Investigator is responsible for ongoing compliance and reporting amendments.

#### Consent to Participate

All participants will provide written informed consent after receiving a clear explanation of the study’s purpose, procedures, risks, and benefits. Participation will be entirely voluntary, and individuals may withdraw at any time without penalty. Personal data will be handled confidentially throughout the study [24, 25].

A copy of the ethics approval and English translation will be submitted with the manuscript.

## Consent for publication

N/A

This study does not include any individual participant data (images, videos, or personal information) that would require consent for publication. All data will be anonymized, and no identifying information will be published. Therefore, consent for publication is not required.

## Authors’ information

The authors represent a multidisciplinary collaboration between academic dentistry, craniofacial research, surgical practice, epidemiology, and pharmaceutical development. Dr. Safdarian, a visiting scholar in the Department of Cleft Lip and Palate at the Arthur A. Dugoni School of Dentistry, University of the Pacific, is engaged in stem cell research using donated dental and soft tissues to explore regenerative solutions for bone and gingival reconstruction in cleft lip and palate patients. The team includes Prof. Amir reza Rokn, Chair of the Implant Department at Tehran University of Medical Sciences; Dr. Amir Jalal Abbasi, Principal Investigator and maxillofacial surgeon; and Prof. Shamshirir, a specialist in epidemiology supporting study design and statistical integrity. Additional collaboration with EmerRx Biopharma contributes to the development of biomaterials for future clinical application. This combined expertise informs a translational, patient-centered approach to the proposed study protocol.

